# Ecological drivers of chikungunya virus transmission: baseline results from a geographically structured, longitudinal study in southern Thailand

**DOI:** 10.64898/2026.02.04.26345533

**Authors:** Erica Rapheal, Darunee Buddhari, Alex Meyer, Taweewun Hunsawong, Sandra Mendoza Guerrero, Stefan Fernandez, Aaron Farmer, Kathryn Anderson, Natalie Dean, T. Alex Perkins, Sarunyou Chusri, Steven T. Stoddard

## Abstract

**Background:** Chikungunya virus (CHIKV) infection can cause significant and long-term morbidity. CHIKV typically appears in explosive outbreaks then vanishes for decades, but evidence from longitudinal studies suggests that it may persist in some populations through low levels of subclinical infection. These epidemiologic dynamics complicate prediction of CHIKV outbreaks and intervention trial planning. Songkhla province in southern Thailand is a promising location for vaccine trials due to its recent history of CHIKV outbreaks (2008 and 2018) and emerging data suggesting low levels of interepidemic transmission.

**Methodology and Principal Findings:** Here, we describe baseline findings from a longitudinal cohort study (2022-2024, n = 5000) of CHIKV transmission in Songkhla. We used serocatalytic models to estimate CHIKV force of infection (FOI), adjusting for risk factors including developed land use surrounding the household, access to garbage collection and clean water, and others. Baseline CHIKV seropositivity in the cohort was 34.6%. Our crude catalytic model estimated 0.0160 (95% CI: 0.0153-0.0168) annual FOI. In adjusted FOI models, a higher proportion of developed land use was associated with an increase in risk of CHIK seropositivity among participants under age 12 (OR: 1.36; 1.25-1.48), who would have been exposed only to the 2018 outbreak, but associated with a decrease in risk (OR: 0.96; 0.94-0.99) among those ages 12 and up, who would have been exposed to two or more outbreaks.

**Conclusions:** These findings suggest that CHIKV infection risk is highly spatially variable, with prediction complicated by historical differences in virus strain and vector. Distinct ecological patterns of exposure are consistent with the 2018-20 outbreak affecting largely urban areas, with little to no exposures in rural areas, while the 2008-09 outbreak was concentrated more heavily in rural areas. Understanding the ecological drivers of this variation has important implications for identifying regions of highest risk for a future CHIKV outbreak.

## Introduction

Chikungunya virus (CHIKV) is a tropical arbovirus spread by the *Aedes* mosquito vectors *Ae. aegypti* and *Ae. albopictus*.^1^ Infection with CHIKV can cause severe sequelae, including debilitating joint pain that may last for years after the acute infection has cleared.^2^ Two vaccines were recently approved in the US and EU based on a surrogate marker of immunity, but CHIKV’s unpredictable epidemiology complicates efficacy study planning and vaccine roll-out strategies.^3–5^ An investigation into adverse effects led to the suspension of one of these vaccine licenses in August, 2025.^6^

Since CHIKV first caused large outbreaks in the Indian Ocean region in 2006, it has expanded globally; today, at least 94 countries or territories have experienced local CHIKV transmission, including France, Italy, and the United States.^7–9^ These outbreaks canonically spread quickly through large populations and are followed by 10-20 years of little to no transmission during the inter-epidemic period. This dynamic is predicted by disease models due to the effect of population immunity on new transmission and is corroborated by the recent reemergence of CHIKV in Guangdong, China and in the French island territory of La Réunion nearly two decades after their last reported outbreaks.^7,10–12^ However, in some regions (most notably Brazil and India), CHIKV is observed every year.^8,9^ Furthermore, several recent longitudinal surveillance studies of chikungunya have found evidence of infection among febrile individuals reporting to surveillance hospitals in non-epidemic years, from 2.2% of acute febrile illnesses in coastal Kenya to 19.3% in southern Thailand.^13,14^

The ecological and epidemiological drivers underlying these seemingly discordant patterns remain underexplored. Few longitudinal cohort studies of CHIKV exist, and even cross-sectional serosurveys, which can provide critical insight into both potential risk factors and local infection history, are uncommon and usually limited in size and scope.^15^ In addition, the similarities in clinical and humoral responses elicited by CHIKV and other arboviruses, especially the dengue (DENV) and Zika (ZIKV) viruses, can make diagnosis challenging and complicate the study of historical surveillance data.

This is particularly problematic in areas with already patchy surveillance and limited capacity for diagnostic testing. Nevertheless, some evidence suggests that a pattern of locally persistent transmission could be driven by ecological variation (i.e., vector-virus interactions) and spatial heterogeneity (i.e., exposure to mosquitoes).^16–19^

The purpose of this study was to evaluate the role of spatial and ecological heterogeneity on current and historic CHIKV transmission patterns, while investigating the persistence, rate, and drivers of interepidemic transmission in southern Thailand. In 2022, we initiated a prospective longitudinal cohort study of CHIKV transmission in the province of Songkhla, where CHIKV has been present for more than 70 years, with two recent outbreaks in 2008/09 and 2018/20.^20,21^ Consistent with the expectation of inter-epidemic lulls in transmission, there were few official reports of CHIKV disease in the years following those outbreaks. However, a febrile illness surveillance study conducted in this area detected cases of CHIKV infection between 2012 and the beginning of the large 2018 outbreak.^14^ This pattern of recurrent outbreaks with evidence of interepidemic transmission suggests that southern Thailand may be uniquely suitable for CHIKV transmission and persistence. Here, we analyze enrollment (baseline) serologic data from our cohort study to describe small-scale spatial variation in the risk of chikungunya infection and disease, characterize patterns in CHIKV transmission history, and identify meaningful risk factors for CHIKV infection in this region.

## Methods

### Study area

This study took place in the southern Thailand province of Songkhla, in the districts of Hat Yai and Na Mom. The study area covers approximately 950 square kilometers (365 square miles), with a total population of more than 400,000. The primary city of Hat Yai is less than 60 km from the border with Malaysia to the south, and 30 km from the Gulf of Thailand to the east.

### Participant recruitment

Participants were recruited using social media, letters, and posters. Interested individuals reached out to study staff and were invited to visit their local health center for enrollment. Study staff also recruited participants actively through visits to the community.

Two analysis cohorts were enrolled: one focused in the dense urban subdistrict of Kho Hong near the enrollment center (“pragmatic” cohort) and another enrolling across a spectrum of ecological contexts, including participants from urban, semi-urban, and rural areas (“ecologic” cohort) (Figure S1). Urban (371-7543/km^2^), semi-urban (129-370/km^2^), and rural (371-7543/km^2^) were defined based on tertiles of subdistrict-level population density. Individuals under age 18 were oversampled (approximately 50% of total enrollees) to improve our power to detect recent outbreaks and estimate an accurate force of infection.

Subjects were considered for inclusion if they met all the following criteria: 1. Healthy, afebrile, and ≥ 2 years of age at the time of enrollment; 2. Written informed consent/assent given; 3. Able to comply with study procedures and available for 3-year study period; 4. No blood or blood transfusion products within the prior 3 months; 5. Not currently immunocompromised or on immunosuppressive therapy.

Up to 10 mL of blood was collected as permitted by weight and age. Participants also completed a short survey, comprising information on demographics (age, sex, etc.), housing infrastructure (roof type, number of windows, etc.), movement and travel history, and more.

Enrollment and follow-up blood samples were tested for CHIKV IgG and IgM using an in-house Enzyme-Linked Immunosorbent Assays (ELISA) with a cutoff of 10 EIA units.^22^

All participants underwent informed consent before enrollment into the study.

### Sample collection and processing

Samples were collected in anticoagulated Vacutainer tubes and processed within 48 hours of collection.

ELISA was used to measure specific antibody response to recombinant E1 (rE1), rE2, and purified inactivated CHIKV whole virus and other arboviruses. This method was modified from Innis et al. (1989) by using specific CHIKV antigen.^23^ Briefly, microtiter plates were sensitized with goat anti-human IgM or IgG antibody and used to capture IgG or IgM antibodies from patient specimens. Viral antigen was then added to the microtiter plate. After incubation, HRP-conjugated secondary antibody was added follow by HRP substrate. The OD450 was measured and calculated for EIA unit. EIA units ≥ 10 were considered CHIKV positive. Here we used the CHIKV rE1 and rE2 assay from Abcam (US).

### Data analysis

Given the history of known CHIKV outbreaks in the region, we divided the dataset into two age strata: individuals under age 12 at enrollment (i.e. born after the 2008-10 outbreak) and those age 12 or higher. Assuming no interepidemic transmission, individuals under age 12 can serve as a proxy for seroincidence in the 2018-20 outbreak. Participants were analyzed in subgroups based on both their enrollment cohort and age strata. Variations in CHIKV seropositivity by specific characteristics were assessed using Pearson’s Chi-squared test (categorical) and Wilcoxon rank sum test (continuous). Seropositivity was calculated both by subgroup and at the subdistrict level. Younger age groups were intentionally oversampled, so the age distribution of the serological sample did not reflect the age structure of the actual population. Overall seroprevalence was estimated by mapping sampled age-specific seropositivities onto the age structure of Thailand in 2022, as reported by the United Nations 2024 World Population Prospects data set.^24^

Attack rate from the outbreak in 2018-20 was calculated using case data from the Thai Ministry of Public Health (MoPH). The total number of cases in each subdistrict reported to MoPH between July 2018 and December 2020 was divided by the subdistrict population. To assess our assumption that seroprevalence among participants under age 12 is a reasonable proxy for seroincidence in the 2018-20 outbreak, we calculated the Pearson’s correlation between these attack rates and our seropositivity under age 12 at the subdistrict level.

ArcGIS Pro 3.4.0 was used to map participant locations and summarize characteristics by subdistrict. Shapefiles were obtained from the Royal Thai Survey Department via the Humanitarian Data Exchange (HDX).^25^ Population density was calculated using 2020 estimates from WorldPop (100m resolution).^26^

To estimate CHIKV force of infection (FOI) – the mean annual risk of infection – we used serocatalytic models with seropositivity to CHIKV as the outcome. We fit binomial models with a *cloglog* link function and a log(age) offset. Models were implemented using the R-INLA package (code adapted from Ribeiro dos Santos et al, 2022).^27^ Model 1 can be seen in the supplement.

We fit crude models for the whole population, as well as populations stratified by age (< 12 and ≥ 12) and by enrollment cohort. To estimate the effect of relevant variables on FOI, we developed saturated models comprising nine hypothesized CHIKV risk factors along with the log(age) offset: developed land use, house type, garbage collection, income, gender, employment, family size, piped water, and air conditioning. These variables were selected to account for indicators of exposure to mosquito biting, proximity to ideal larval development areas, and measures of socioeconomic status. Specific details of how these variables are defined can be found in the supplement. Saturated models were then reduced to only relevant variables using a 10% reverse selection method, in which variables are removed from the model sequentially. The variable with the least impact on the estimate of FOI is dropped and the process is repeated until no variable can be removed without changing the FOI at least 10%. Both the saturated and reduced models were also stratified by under/over age 12 and enrollment cohort.

All analyses were performed using R-4.3.1 and ArcGIS Pro 3.4.0.

### Ethical considerations

This study was approved by the Walter Reed Army Institute of Research Institutional Review Board (Silver Spring, MD, USA) and the Office of Human Research Ethics Committee at Prince of Songkhla University (Hat Yai, Songkhla, Thailand).

## Results

### Study population

Between 2022 and 2024, 5000 individuals were enrolled in this study. Of these, 4994 reside within the study area and were included in this analysis. A total of 2195 participants (44%) were under age 18 at enrollment, and 1471 (29%) were under age 12. The pragmatic cohort contained 2494 participants, of which 634 were under age 18 (25%) and 366 were under age 12 (12%). The ecologic cohort contained 2500 participants, with 1556 under age 18 and 1102 (62%) under age 12 (44%).

Participants were an average of 30 years old (median 25, ranging from 2 to 88 years) at enrollment. 60% of enrollees were female (Table 1). More than half of all participants (n = 3119) were recruited from highly dense areas in the city of Hat Yai; the remainder were distributed throughout the study area (Figure 1). Age distribution in the study population and in the underlying population can be seen in Figure 2.

**Figure 1.**
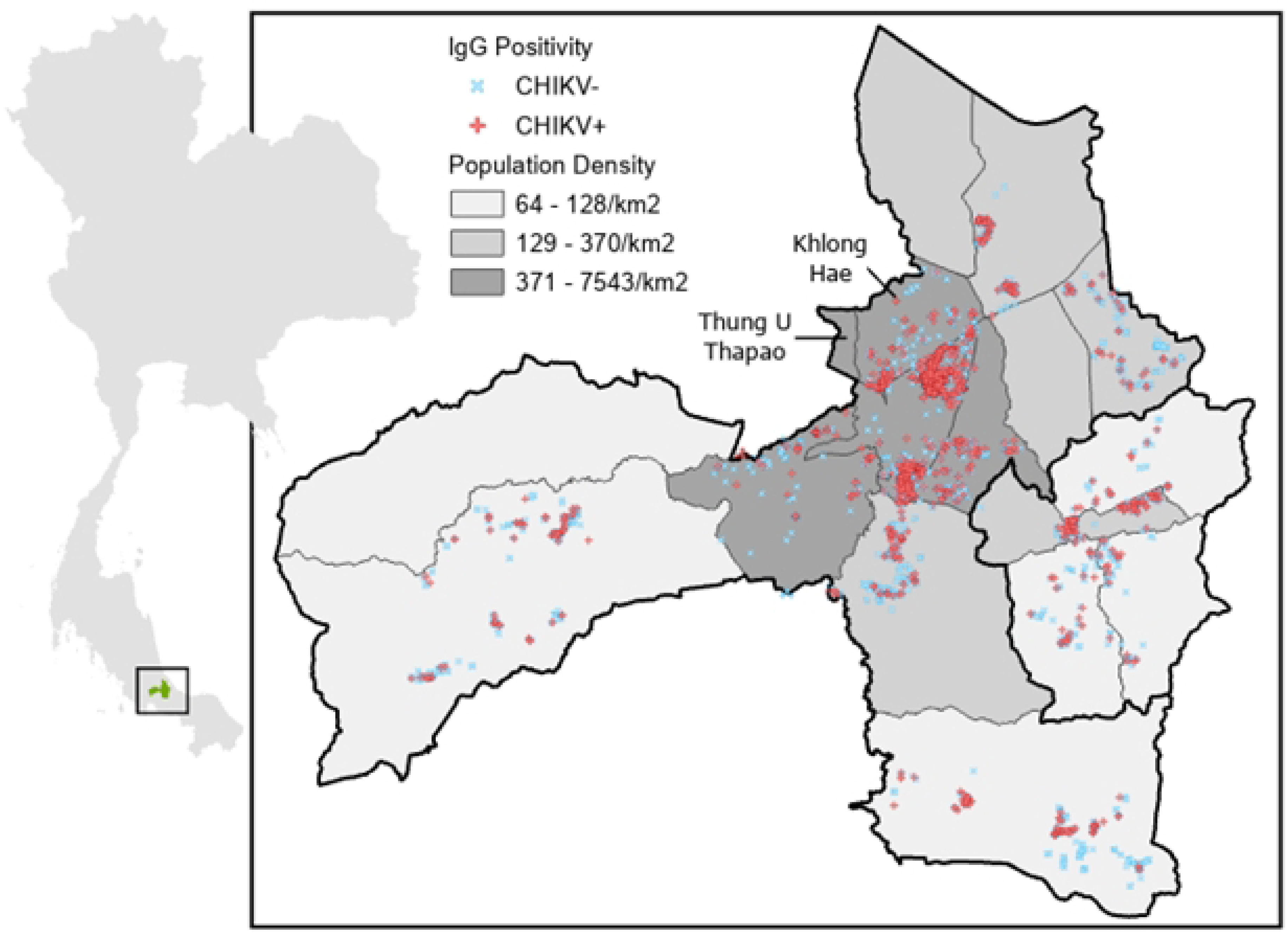
Map of Hat Yai area with study participants. Map shows 17 subdistricts in the districts of Hat Yai and Na Mom in Songkhla province, Thailand. Points show participant households. Red points indicate CHIKV+, while blue points indicate CHIKV-. Population density is displayed in tertiles at the subdistrict level; rural: 64-128/km^2^, semi-urban: 129-370/km^2^, urban: 371-7543/km^2^. Districts of Hat Yai (north) and Na Mom (south) are indicated with thick black borders.

**Figure 2.**
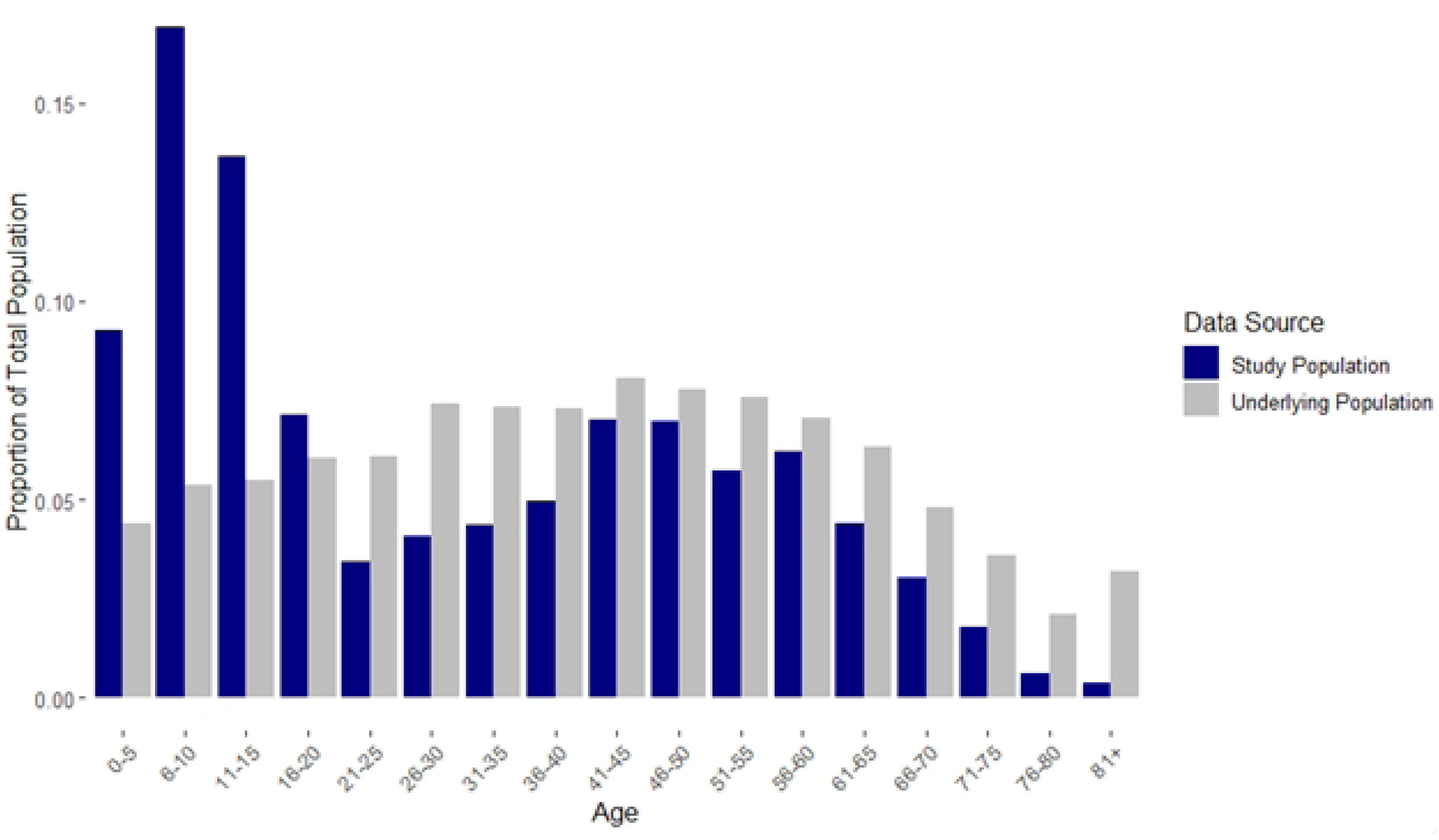
Age distribution of underlying population and study population. Age distribution in 5-year intervals for the full study population (blue, n = 4994) and for the underlying population (gray). Age distribution in the underlying population was calculated using publicly available Thai census data.^29^

**Table 1.** Population characteristics.

| Characteristic | Serostatus |  |  | p-value |
| --- | --- | --- | --- | --- |
|  | Overall<br>N = 4994 | CHIKV-<br>N = 3254 | CHIKV+<br>N = 1740 |  |
| Age | 25 (10, 48) | 14 (8, 40) | 45 (23, 58) | <0.001 |
| Gender |  |  |  | <0.001 |
| Female | 2,981 (60%) | 1,852 (57%) | 1,129 (65%) |  |
| Male | 2,013 (40%) | 1,402 (43%) | 611 (35%) |  |
| Employment |  |  |  | <0.001 |
| Not Employed | 1,211 (24%) | 643 (20%) | 568 (33%) |  |
| Employed/Student | 3,783 (76%) | 2,611 (80%) | 1,172 (67%) |  |
| House type |  |  |  | <0.001 |
| Single House | 2,894 (58%) | 1,953 (60%) | 941 (54%) |  |
| Townhouse/Apartment | 1,948 (39%) | 1,190 (37%) | 758 (44%) |  |
| Standing on Poles | 152 (3.0%) | 111 (3.4%) | 41 (2.4%) |  |
| Garbage collection |  |  |  | <0.001 |
| No | 1,114 (22%) | 790 (24%) | 324 (19%) |  |
| Yes | 3,880 (78%) | 2,464 (76%) | 1,416 (81%) |  |
| Piped water |  |  |  | 0.36 |
| No | 1,813 (36%) | 1,196 (37%) | 617 (35%) |  |
| Yes | 3,181 (64%) | 2,058 (63%) | 1,123 (65%) |  |
| Air conditioning |  |  |  | 0.094 |
| No | 3,382 (68%) | 2,230 (69%) | 1,152 (66%) |  |
| Yes | 1,612 (32%) | 1,024 (31%) | 588 (34%) |  |
| Population density |  |  |  | <0.001 |
| Rural | 808 (16%) | 606 (19%) | 202 (12%) |  |
| Suburban | 1,067 (21%) | 805 (25%) | 262 (15%) |  |
| Urban | 3,119 (62%) | 1,843 (57%) | 1,276 (73%) |  |
| Monthly income (1,000 baht, IQR) | 20 (10, 30) | 20 (10, 30) | 18 (10, 30) | 0.013 |
| Family size (count, IQR) | 4 (3, 6) | 4 (3, 6) | 4 (3, 5) | <0.001 |
| Land use (% of 500m surrounding household, IQR) |  |  |  |  |
| Developed | 95 (64, 100) | 91 (57, 100) | 98 (78, 100) | <0.001 |
| Agriculture/Short vegetation | 0.9 (0.0, 4.4) | 1.2 (0.1, 5.4) | 0.3 (0.0, 2.5) | <0.001 |
| Forest | 2 (0, 23) | 4 (0, 28) | 0 (0, 10) | <0.001 |

Continuous variables: median (interquartile range), p-values calculated using Wilcoxon rank sum test. Categorical variables: n (%), p-values calculated using Pearson's Chi-squared test. Excludes individuals who did not reside in the study area.

### Seropositivity for anti-CHIKV IgG antibodies

We found a mean anti-CHIKV IgG seropositivity rate of 34.8% (1740/4994) in our cohort. After adjustment for underlying age structure, we estimated IgG seroprevalence of 43.6% (95% credible interval: 42.1-44.7) for the full population and 47.3% (46.1-52.4) among those over age 12.

In unadjusted analyses (Pearson’s Chi-square/Wilcoxon rank sum test), having a higher proportion of developed land use in the 500m surrounding the household was significantly associated with increased risk of CHIKV seropositivity, along with increasing age, female gender, not being employed/student, living in a townhouse or apartment, having access to garbage collection, living in a rural subdistrict, lower monthly income, and smaller family size (Table 1).

### Force of infection models

Our crude catalytic model estimated 0.0160 (95% CI: 0.0153-0.0168) annual FOI for the whole population. Among individuals under 12, this estimate was 0.0155 (95% CI: 0.0133-0.0182); among 12 and over it was 0.0160 (95% CI: 0.0152-0.0169).

In saturated FOI models, different variables were predictive of seropositivity among different age groups and cohorts (Table 2). In the full population (all ages and cohorts), only garbage collection and family size were retained in the reduced model, and only family size was a significant predictor of CHIKV seropositivity (OR: 1.05; 95% CI: 1.02-1.08).

**Table 2.**
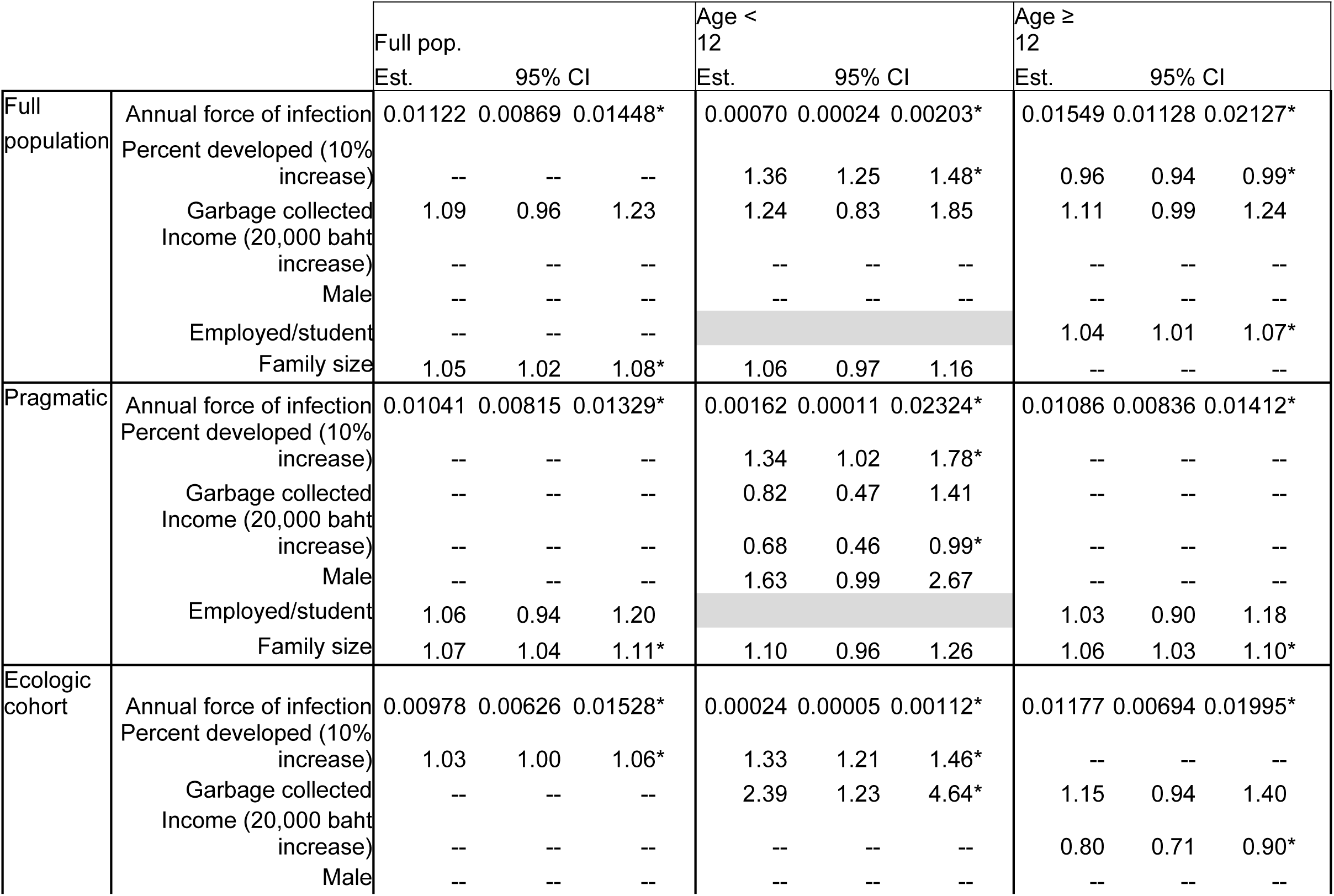
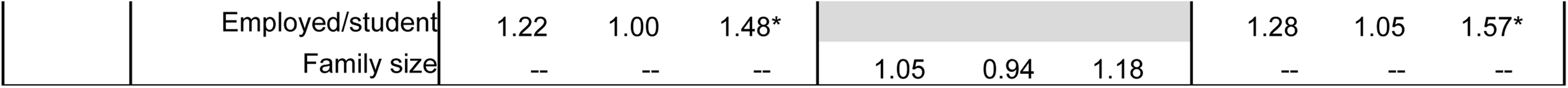
Reduced catalytic model outputs stratified by age and enrollment cohort.

Output from reduced catalytic models among nine population subsets. All models started with ten hypothesized variables. Models were reduced using 10% reverse model selection, in which variables are successively removed from the model. Variables that change the estimate for force of infection by less than 10% are removed from the model. A blank cell indicates that that variable was not retained in the final model. Cells with gray shading were not considered for that population. Living in an apartment/townhouse, access to piped water, air conditioning, and the income by employment interaction were also considered in these models but were not retained in any model. Estimates are odds ratios except force of infection. Percent developed estimates OR for a 10% increase in developed land use in a 500m radius around participant home. Income estimates OR for a 20,000 baht increase in monthly income. *p-value ≤ 0.05.

When this population was stratified by age, differences in the impact of ecological and environmental factors emerged; among participants under age 12, an increase in developed land use was associated with an increase in CHIK positivity (OR: 1.36; 1.25-1.48). However, among those ages 12 and up, the same increase in developed land use was associated with a significant decrease in CHIK positivity (OR: 0.96; 0.94-0.99).

As these age strata were further stratified into pragmatic (urban) and ecologic (urban, semi-urban, and rural) cohorts, more patterns arise. In the pragmatic cohort under 12 group, developed land use, garbage collection, income, gender, and family size were retained in the reduced model. For the same age group but in the ecologic cohort, only development, garbage collection, and family size were retained. In both under 12 models, developed land use was significantly associated with the outcome (OR of approximately 1.33 in both models). Garbage collection was significantly positively associated with seropositivity in the ecologic cohort (OR: 2.39; 1.23-4.64) but negatively (though not significantly) associated in the pragmatic cohort (OR: 0.82; 0.47-1.41) (Table 2).

Estimated FOI in the reduced models was lowest among participants under 12 in the ecologic cohort (0.00024 mean annual FOI) and highest among those ages 12 and over in the full population (0.01549 mean annual FOI).

### Spatial variation in seropositivity

Average subdistrict-level CHIKV seropositivity in our study population ranged from 15% in semiurban Tha Kham to 42% in urban Kho Hong. In general, seroprevalence was higher in urban subdistricts (34-42%) and lower in rural and semiurban subdistricts (15-29%) (Table 3).

**Table 3.** Enrollment and CHIKV seropositivity by subdistrict in cohort.

| Subdistrict | Total Population | Population density | N |  |  | CHIKV seropositivity |  |  |
| --- | --- | --- | --- | --- | --- | --- | --- | --- |
|  |  |  | Total | Age < 12 | Age ≥ 12 | Total | Age < 12 | Age ≥ 12 |
| *Total* |  |  | 4994 | 1539 | 3455 | 35% | 11% | 45% |
| Hat Yai | 140,579 | Urban | 167 | 41 | 126 | 34% | 15% | 40% |
| Khlong Hae | 44,816 | Urban | 287 | 151 | 136 | 37% | 24% | 52% |
| Kho Hong | 46,169 | Urban | 2,494 | 382 | 2,112 | 42% | 18% | 46% |
| Khuan Lang | 52,919 | Urban | 171 | 50 | 121 | 37% | 18% | 45% |
| Ban Phru | 36,759 | Semi-urban | 255 | 134 | 121 | 24% | 13% | 36% |
| Na Mom | 8,415 | Semi-urban | 329 | 110 | 219 | 28% | 2% | 41% |
| Nam Noi | 15,501 | Semi-urban | 231 | 105 | 126 | 29% | 12% | 42% |
| Tha Kham | 9,791 | Semi-urban | 252 | 147 | 105 | 15% | 3% | 32% |
| Khlong Rang | 4,359 | Rural | 72 | 37 | 35 | 25% | 0% | 51% |
| Pathong | 13,598 | Rural | 287 | 151 | 136 | 25% | 3% | 50% |
| Phichit | 4,658 | Rural | 52 | 27 | 25 | 29% | 0% | 60% |
| Thung Khamin | 5,634 | Rural | 105 | 47 | 58 | 25% | 0% | 45% |
| Thung Tam Sao | 17,403 | Rural | 292 | 157 | 135 | 24% | 4% | 47% |

Subdistrict population was determined using publicly available Thai census data.^29^ Population density is displayed in tertiles at the subdistrict level; rural: 64-128/km^2^, semi-urban: 129-370/km^2^, urban: 371-7543/km^2^. CHIKV seropositivity shows the percentage of individuals in the cohort who were positive for anti-CHIKV IgG.

However, these patterns in seropositivity may have been driven by differences in age as well as environment. Age stratification again shows that spatial patterns of exposure differed significantly between the outbreaks in 2008-9 and 2018-19 (Figure 3a), with those exposed to only the 2018 outbreak being mostly urban and those exposed to two or more outbreaks more rural. Among participants in both cohorts who were under age 12 at enrollment (born after 2010), CHIKV seroprevalence was highest in urban subdistricts near the city of Hat Yai (15-24%). In three rural subdistricts (Khlong Rang, Phichit, and Thung Khamin), there were no participants under the age of 12 who were CHIKV positive. Among adults and children over 12 years, however, seroprevalence was highest (45-60%) in rural areas, and lowest (32-42%) in semiurban areas (Table 3).

**Figure 3.**
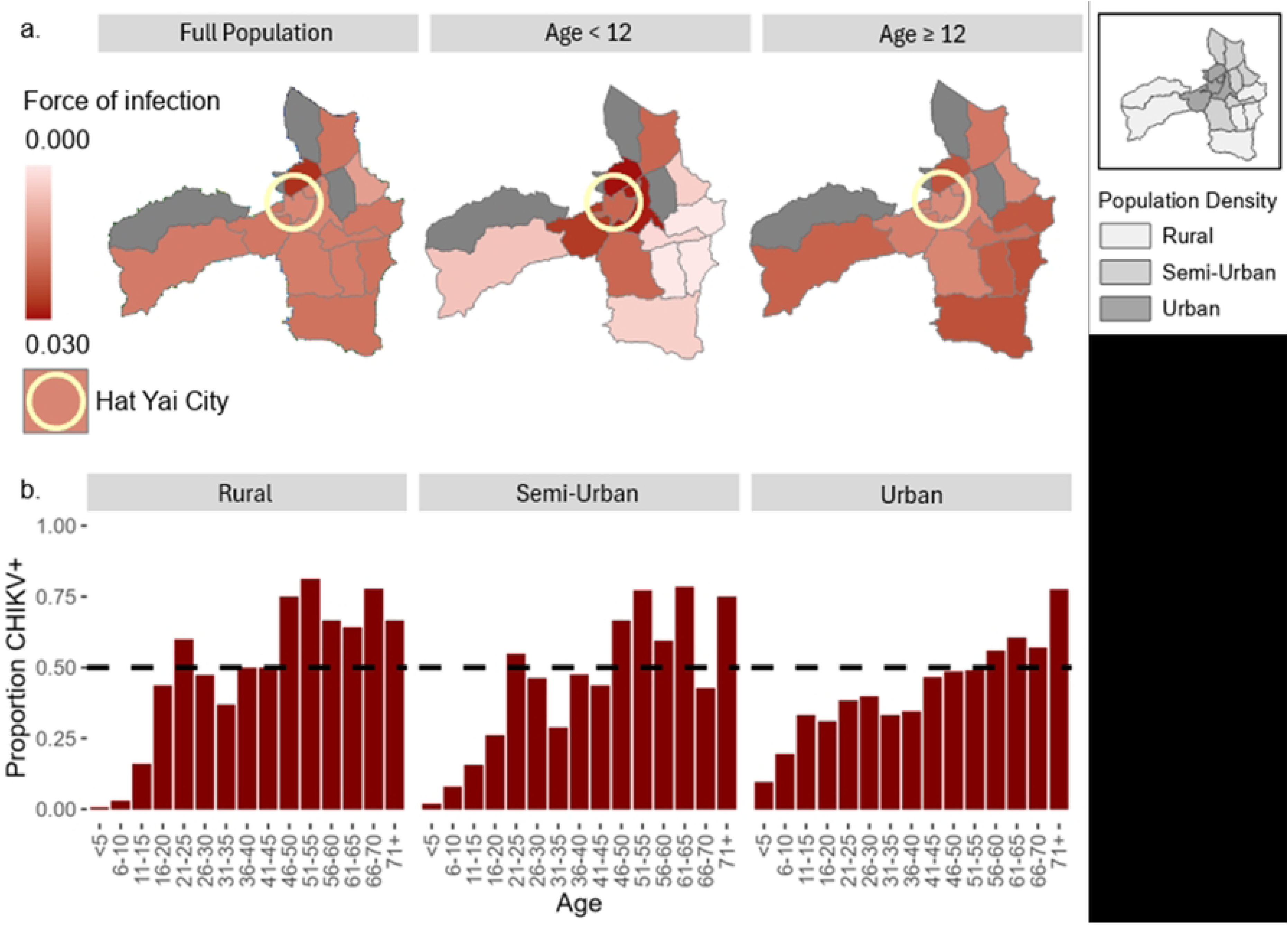
Force of infection among study participants stratified by age and subdistrict. Crude force of infection by subdistrict among full population, < age 12, and ≥ age 12. Population density is provided in the inset for reference. b. CHIKV seropositivity by age in rural, semiurban, and urban subdistricts. Dotted line indicates 50% seropositivity.

Estimates of FOI also varied spatially, from 0.011 in Tha Kham (semi-urban) to 0.027 in Khlong Hae (urban). Subdistrict-level differences in FOI between age strata can be seen clearly in Figure 3: among participants under 12, the crude FOI was highest in urban areas; among those over 12, it was highest in rural areas. Children under 12 in rural areas again showed almost no evidence of CHIKV transmission.

### Case reports from Thai MoPH

A total of 2723 CHIKV disease cases were reported to the Thai MoPH between July 20, 2018 and December 26, 2020.^28^ Most of these cases were reported in 2018 (n = 1627) and 2019 (n = 1024). The attack rate during the full 29-month period was 0.67%, or 67/10,000. Subdistrict-level case attack rates from the 2018-20 outbreak were highest in the urban subdistricts of Hat Yai (0.89%), Kho Hong (0.80%), and Khuan Lang (0.70%). These attack rates were highly correlated with subdistrict-level seroprevalence among study participants under age 12 (R = 0.75, p = 0.0031; Figure 4).

**Figure 4.**
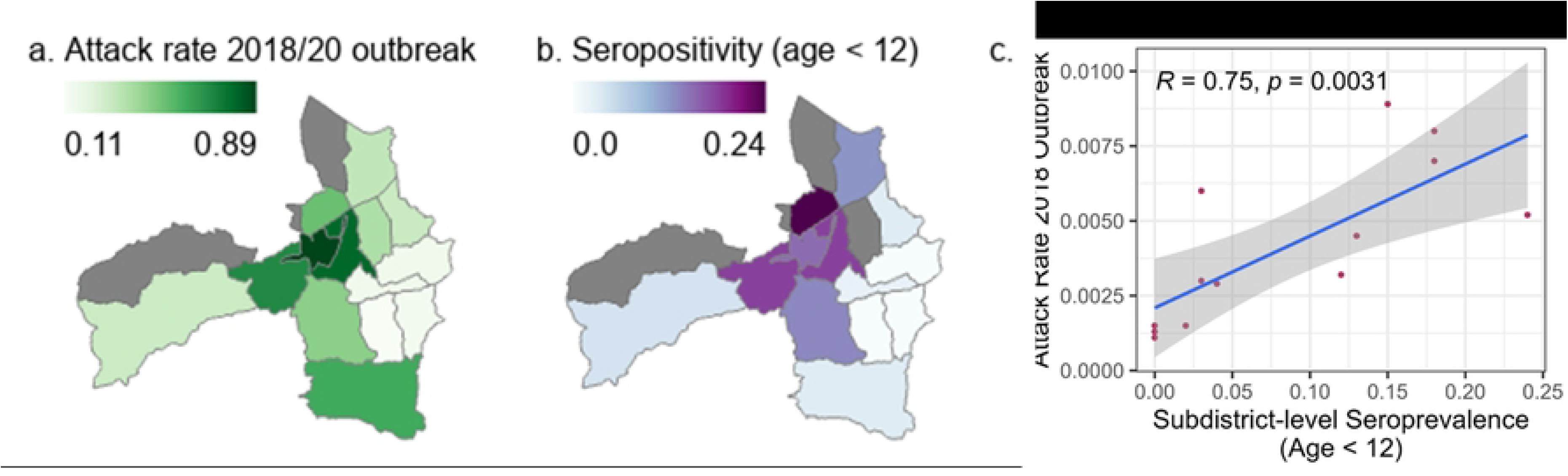
Correlation between 2018-20 attack rate and seropositivity at the subdistrict level. a. Attack rate from 2018-20 CHIKV outbreak calculated from total cases reported to the Ministry of Public Health and subdistrict population. b. Subdistrict-level seroprevalence in study population under age 12. c. Pearson’s correlation between subdistrict-level attack rate in 2018/19 and subdistrict-level seroprevalence (R = 0.75, p = 0.0031).

## Discussion

Baseline serological samples collected in southern Thailand indicate that CHIKV seropositivity in this region is spatially heterogeneous and varies across a gradient of urbanicity. Distinct ecological patterns of exposure are consistent with the 2018-20 outbreak affecting largely urban areas, with little to no exposures in rural areas, while the 2008-09 outbreak was concentrated more heavily in rural areas. When models were stratified by age and enrollment cohort, different and sometimes contradictory sets of variables were retained. Such discrepancies suggest a complex and multifaceted relationship between ecology, demographic characteristics, and risk of CHIKV infection.

We found an overall CHIKV seropositivity of 34.8% in the cohort, corresponding with 43.6% in the population after adjusting for differences in cohort and population age structure. This is slightly higher than previously reported estimates of seroprevalence – approximately 27% in Thailand and 6% in Malaysia – though these estimates were made before the outbreak in 2018.^15,30,31^ Our adjusted FOI in the reduced, full population model was 0.011. This is consistent with previously modeled estimates from southern Thailand and the Malay Peninsula, which fall between around 0.01 and 0.02.^9,15^ Variation in CHIKV FOI across an ecological spectrum at this small scale has not previously been published to our knowledge, so it is difficult to assess whether our findings are typical of CHIKV transmission patterns. Reports of CHIKV incidence at the municipality level in Brazil show significant heterogeneity in transmission at a small spatial scale, but these findings were not explicitly linked to ecology, nor did they include serologic data.^17^

In our reduced FOI models, few variables were retained consistently. Employment/student had a positive impact on odds of seropositivity, particularly in the ecologic cohort, in which it was associated with a 22% (95% CI: 0-48%) increase in odds. This association suggests that time spent at home may be protective against infection with CHIKV, at least in the ecologically diverse population. That the employed seem to be at higher risk may be related to the increased risk of exposure associated with rubber plantation farming, which is a primary occupation in rural parts of this region. Analysis of more in-depth movement and occupation data would help to elucidate this finding. Interestingly, measures of socioeconomic status, including income, piped water, air conditioning, and garbage collection, had limited significance in this population. None of these was significant in the full population model. Garbage collection was retained in six of the nine models and in all cases was positively correlated with seropositivity, which contradicts the expectation that lack of garbage infrastructure would increase exposure to mosquitoes due to higher density of breeding sites. However, garbage collection may be more common in urban areas, which suggests that the strong association between developed land use and seropositivity may be helping to drive the association with garbage collection.

Across subdistricts in our study, cohort seroprevalence varied from 15-40%, exhibiting a spatial heterogeneity in past transmission patterns that is consistent with reports from Brazil.^17^ These patterns were noticeably different among those born before 2010, who would have been exposed to at least two CHIKV outbreaks, and those born in or after 2010, who would have experienced only the 2018/19 outbreak. Though we were unable to directly address reports of ongoing interepidemic transmission in this baseline analysis, our findings suggest that there was very limited transmission in rural areas both during the 2010-18 interepidemic period and during the 2018-20 outbreak. In our study population, three rural subdistricts had zero seropositive individuals under age 12, and in the other two rural subdistricts less than 5% of the participants were seropositive. The seroprevalence estimates among ages 12 and up in these subdistricts, meanwhile, were some of the highest in the study area, ranging from 45-60%. In addition, in the stratified models, developed land use was associated with an increase in risk among ages under 12 and a decrease in risk among ages 12 and over. Case data from the Thai MoPH and the Malaysian Ministry of Health, also show that cases reported during the 2018 outbreak were concentrated in urban areas, while prior transmission was focused in more rural populations.^28,32–34^

However, previously reported evidence of interepidemic transmission from a nearby hospital contradicts these findings; Farmer et al found that 19% of febrile illness cases reporting to the hospital during the interepidemic period (2012-2017) were attributable to CHIKV infection, suggesting ongoing transmission of CHIKV.^14^ This contradiction perhaps suggests that the exposure profile among children is not representative of rural populations as a whole, despite the strong correlation between seroprevalence in children under 12 and case attack rate.^14^ This may be related to the finding noted above, that employed people (likely rubber plantation workers) had increased risk of seropositivity. While the Famer study did not explicitly distinguish participants living in urban or rural areas, they did identify more than 300 cases of chikungunya among agricultural workers, who are likely to live in more rural areas.

It is not clear which factors may have contributed to this sustained transmission between 2012 and 2017, particularly considering the lack of evidence of transmission since 2010 in rural areas in our data. Rural villages are spatially dispersed in the region around Hat Yai which may have led to local, small-scale outbreaks sustained by regional human movements and patchy population immunity during the period. In addition, while affected rural populations would have had high seroprevalence after the outbreak ended in 2009, semi-urban and urban populations did not, potentially providing susceptible hosts for sustained transmission in the region.

Spatial heterogeneity in seroprevalence and transmission patterns over relatively fine spatial scales could be driven by host and vector mobility and variation in local conditions at the time of an outbreak. Clustering, combined with the effects of rapidly increasing local population immunity and changing (seasonal) weather conditions, could lead to virus fade-out before the virus can spread fully through a population and would result in a patchy pattern of seroprevalence.^16,17^ In the case of CHIKV, the existence of distinct strains, each adapted to one of two co-circulating mosquito vectors, also complicates the epidemiologic landscape. Azman et al (2024) have previously shown that CHIKV transmission in Malaysia in 2008 was driven largely by *Ae. albopictus,* which is more abundant in rural areas, while the 2018 outbreak was driven by *Ae. aegypti,* which prefers more densely populated areas.^32,35^ Indeed, CHIKV strains isolated in the Hat Yai region prior to the 2018 outbreak had the E1-226A mutation favoring *Ae. albopictus* while the strain that caused the 2018 outbreak had mutations favoring *Ae. aegypti*.^36^ Thus, a vector by virus interaction in regions where both *Aedes* mosquito species are present could also contribute to spatial variation in transmission and patterns of seroprevalence.

This project had several limitations. Importantly – particularly in the context of discrete, epidemic transmission patterns – the catalytic models used here assume a constant FOI. If, as found by Farmer et al, there is consistent, low-level interepidemic transmission, this assumption may be close to correct; though the FOI in epidemic years is necessarily larger than non-epidemic, these years are relatively rare. Though the constant FOI has been used to effectively model CHIKV in other published analyses, future work may consider more complex models.^15^ In addition, we have focused here on developed land use (on the individual level) and population density (at the subdistrict level) as metrics for urbanicity. While we identified clear patterns in seropositivity using these variables, urbanicity is a nebulous concept that has been defined inconsistently. Different metrics may predict different elements of transmission risk. We chose land use at a small (500m) radius here due to our interest in vector dynamics. A better understanding of the elements of urbanicity and how they impact infection risk would help to further clarify these associations. Finally, we know very little about the actual distribution of vectors in this population. While we can use our understanding of both vector preferences and availability of mosquito breeding sites (as estimated by access to garbage collection, running water, etc.) to make assumptions about which mosquitos are active in which areas, without species-specific mosquito studies, we lack explicit confirmation for our hypotheses about vector dynamics.

Our findings show that urbanicity and related features have a significant impact on CHIKV transmission heterogeneity. The specific mechanism by which this occurs is likely a combination of human movement patterns, historical seroprevalence, and density of mosquito breeding and biting sites. These drivers of CHIKV transmission are patchy and vary across an ecological gradient, which results in variation in seropositivity at a small spatial scale. As longitudinal data from this study are analyzed, we may be able to identify changes in serology over time, which would help elucidate the relative contributions of these risk factors, while helping to guide vaccine development and deployment and more accurately model infection risk. This project represents an important advancement in our understanding of CHIKV transmission and ecology in Thailand, demonstrating that CHIKV and other *Aedes*-borne viruses have a complex and dynamic relationship with human population structure, vector ecology, and viral evolution.

## Data Availability

Data are available upon request and ethical approval. This study uses human subjects data from an active protocol, and data can therefore not be shared publicly.

## Acknowledgements

We would like to thank our study participants for their willingness to participate in this project, the research team at PSU and AFRIMS, including Dr. Edgie Mark Co, for their work in completing enrollment and lab work, Kelly Warfield and Sarah Royalty-Tredo for their support during study setup, and MIDRP (MI210177) and Emergent BioSolutions/Bavarian Nordic Inc for funding this project.

Material has been reviewed by the Walter Reed Army Institute of Research. There is no objection to its presentation and/or publication. The opinions or assertions contained herein are the private views of the author, and are not to be construed as official, or as reflecting true views of the Department of the Army or the Department of Defense. The investigators have adhered to the policies for protection of human subjects as prescribed in AR 70–25.

